# The Efficacy of Facemasks in the Prevention of COVID-19: A Systematic Review

**DOI:** 10.1101/2022.07.28.22278153

**Authors:** Bedir Alihsan, Arrianna Mohammed, Yash Bisen, Janice Lester, Christian Nouryan, Joseph Cervia

## Abstract

Facemasks have become a symbol of disease prevention in the context of COVID-19; yet, there still exists a paucity of collected scientific evidence surrounding their epidemiological efficacy in the prevention of SARS-CoV-2 transmission. This systematic review sought to analyze the efficacy of facemasks, regardless of type, on the prevention of SARS-CoV-2 transmission in both healthcare and community settings.

The initial review yielded 1732 studies, which were reviewed by three study team members. Sixty-one full text studies were found to meet entry criteria, and 13 studies yielded data that was used in the final analysis.

In all, 243 subjects were infected with COVID-19, of whom 97 had been wearing masks and 146 had not. The probability of getting COVID-19 for mask wearers was 7% (97/1463, p=0.002), for non-mask wearers, probability was 52% (158/303, p=0.94). The Relative Risk of getting COVID-19 for mask wearers was 0.13 (95% CI: 0.10-0.16).

Based on these results, we determined that across healthcare and community settings, those who wore masks were less likely to contact COVID-19. Future investigations are warranted as more information becomes available.

## Introduction

Severe acute respiratory syndrome coronavirus 2 (SARS-CoV-2), a member of the Coronaviridae family, is the causative agent of COVID-19. It is the seventh known coronavirus to infect humans: others include SARS-CoV-1, the causative agent of SARS, MERS-CoV, the causative agent of MERS, as well as HCoV-229E, -NL63, -OC43, and -HKU1 which are endemic human coronaviruses and are among the causative agents of the common cold.^1^ SARS-CoV-2 is reported to have emerged from the Huanan South China Seafood Market in Wuhan City, Hubei Province, China in late 2019. The virus quickly spread throughout the country, and then across the world. In response, the WHO declared a global health emergency on January 31, 2020, and later a pandemic on March 11, 2020.^2^

Despite the swiftness with which the virus spread, responses to the pandemic varied greatly by country. Throughout the course of the pandemic, the use of facemasks in prevention of the transmission of SARS-CoV-2 has remained one of the most contentious topics. Many East Asian countries, who had experience with the SARS epidemic in the early 2000s were quick to recommend that their citizens wear facemasks in public. Many of these countries had a public which remembered the SARS epidemic and had experience using facemasks in the prevention of viral transmission.^3^ The Chinese government issued guidelines recommending the personal use of facemasks on January 31, 2020.^4^ The government of Hong Kong recommended that its citizens use facemasks as early as January 24, 2020.^5^ In contrast to this, the CDC did not recommend that U.S. citizens wear facemasks until early April^6,^ and the WHO did not officially recommend that members of the public wear masks until June 5, 2020.^7^

At the beginning of the pandemic, disruption of supply chains and increased use of personal protective equipment (PPE) caused concerns of widespread shortages, including facemasks.^8^ These concerns may have influenced some of the initial recommendations by the CDC and WHO, as officials feared that widespread use of facemasks might exacerbate PPE shortages, thus limiting supplies for healthcare workers treating COVID-19 in hospital settings. These initial conflicting guidelines by government and international bodies became a source of confusion in the general public, and contributed to the politicization of facemask use in places like the U.S.^9^

Despite disparity in facemask use by country early in the pandemic, many public policy decisions were made in the absence of guidance from peer-reviewed scientific sources. Even though the facemask has become a symbol of disease prevention in the context of COVID-19, there still exists a paucity of collected scientific evidence surrounding the epidemiological efficacy of facemasks in the prevention of SARS-CoV-2 transmission. This systematic review sought to analyze the efficacy of facemasks, regardless of type, on the prevention of SARS-CoV-2 transmission in both healthcare and community settings. It was hypothesized that wearing a facemask would be associated with lower rates of COVID-19.

## Methods

A systematic review was conducted to identify relevant studies. A medical librarian conducted a literature search utilizing Pubmed, Web of Science, Embase and Cochrane library from April 2020 to August 2020. Citations were deduplicated using Covidence.org. Only English language articles were retrieved, and conference proceedings were omitted. Results were reported according to the Preferred Reporting Items for Systematic Reviews and Meta-Analyses (PRISMA) criteria. The following broad strategy was utilized:

> Masks: Mask[text word(tw)] OR masks[tw] OR facemask[tw] OR facemasks[tw] OR “face mask”[tw] OR “face masks”[tw] OR “face covering”[tw] OR “face coverings”[tw] OR “masks[medical subject headings (mesh)].”
>
> COVID-19: “COVID-19”[Supplementary Concept] OR “severe acute respiratory syndrome coronavirus 2”[Supplementary Concept] OR “Coronavirus Infections”[Mesh] OR “COVID-19”[tw] OR “covid19”[tw] OR “covid2019”[tw] OR “ncov2019”[tw] OR “ncov-2019”[tw] OR “2019-nCoV”[tw] OR “2019nCoV”[tw] OR “nCoV”[tw] OR “2019 ncov”[tw] OR “2019nCoV”[tw] OR “COV 2”[tw] OR “CoV2”[tw] OR “SARS-CoV-2”[tw] OR “SARSCoV2”[tw] OR “sars cov 2”[tw] OR “sars coronavirus 2”[tw] OR “HCoV-19”[tw] OR “novel coronavirus”[tw] or “covid”[tw] OR “coronavirus disease 2019”[tw] OR 2019 “novel coronavirus disease”[tw] OR “covid-19 pandemic”[tw] OR “2019 novel coronavirus infection”[tw] OR “SARS-CoV-2 infection”[tw] or “2019-nCoV infection”[tw] or “COVID-19 virus disease”[tw] or “2019 novel coronavirus infection”[tw] or “2019-nCoV disease”[tw].

The initial review yielded 1732 studies, which were reviewed by three study team members. Sixty-one full text studies were found to meet the criteria, and 13 studies were used in the final analysis. (Figure 1) Frequencies, relative risk, confidence intervals and t-tests were calculated where appropriate, to measure differences between groups who reported wearing masks vs. not wearing masks for the overall study group, as well as health care, and community settings.

**Figure 1.**
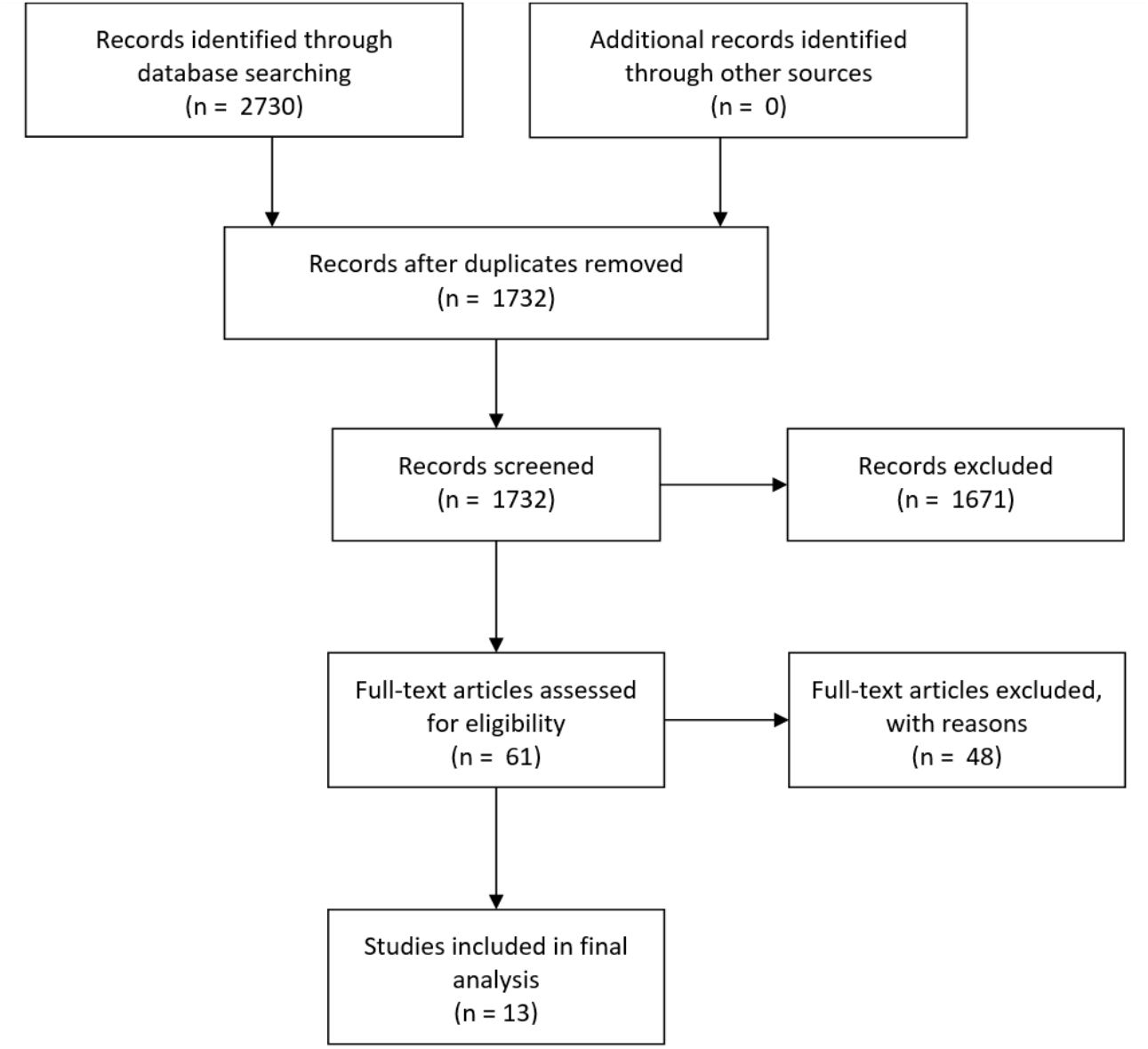
Flow chart of systematic review.

## Results

*Figure 2* - We analyzed 1539 overall subjects from 13 studies (4 community-based, 9 health care-based, Table 1) which reported the full amount of data required. In all, 243 subjects were infected with COVID-19, of whom 97 had been wearing masks and 146 had not. The probability of getting COVID-19 for mask wearers was 7% (97/1463, p=0.002), for non-mask wearers, probability was 52% (158/303, p=0.94). Relative Risk of getting COVID-19 for mask wearers was 0.13 (95% CI: 0.10-0.16).

**Figure 2.**
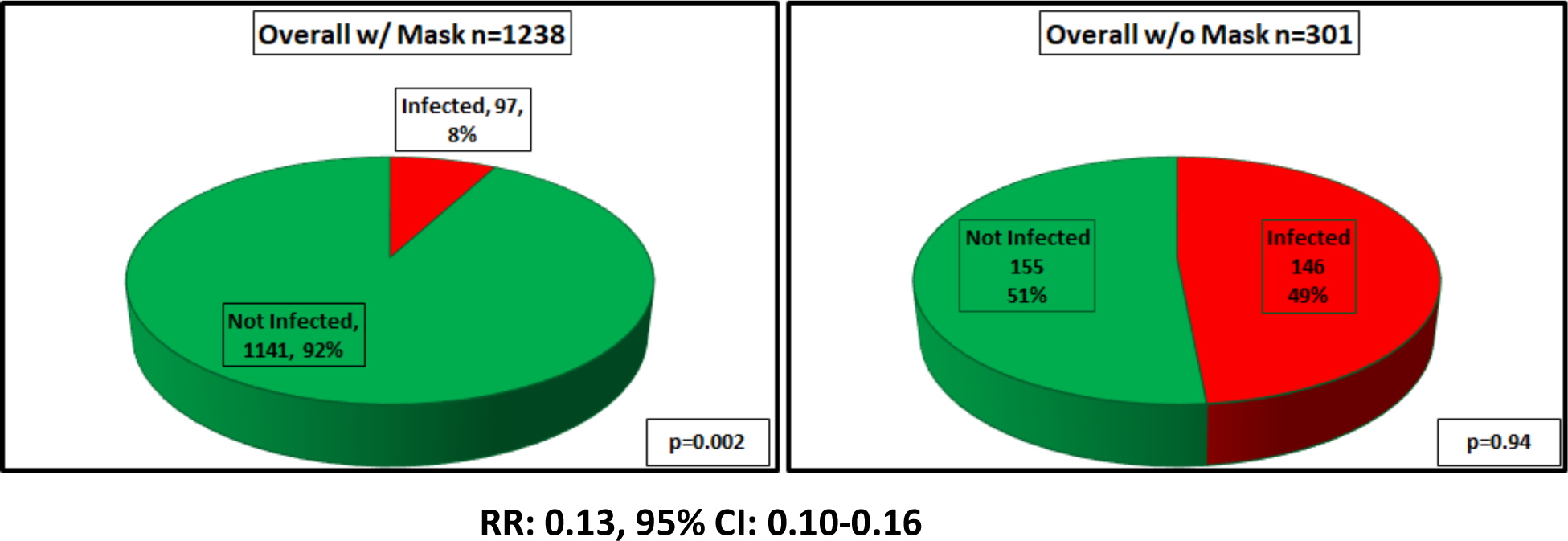
Total Subjects Infected and Not Infected with COVID-19.

**Table 1.**
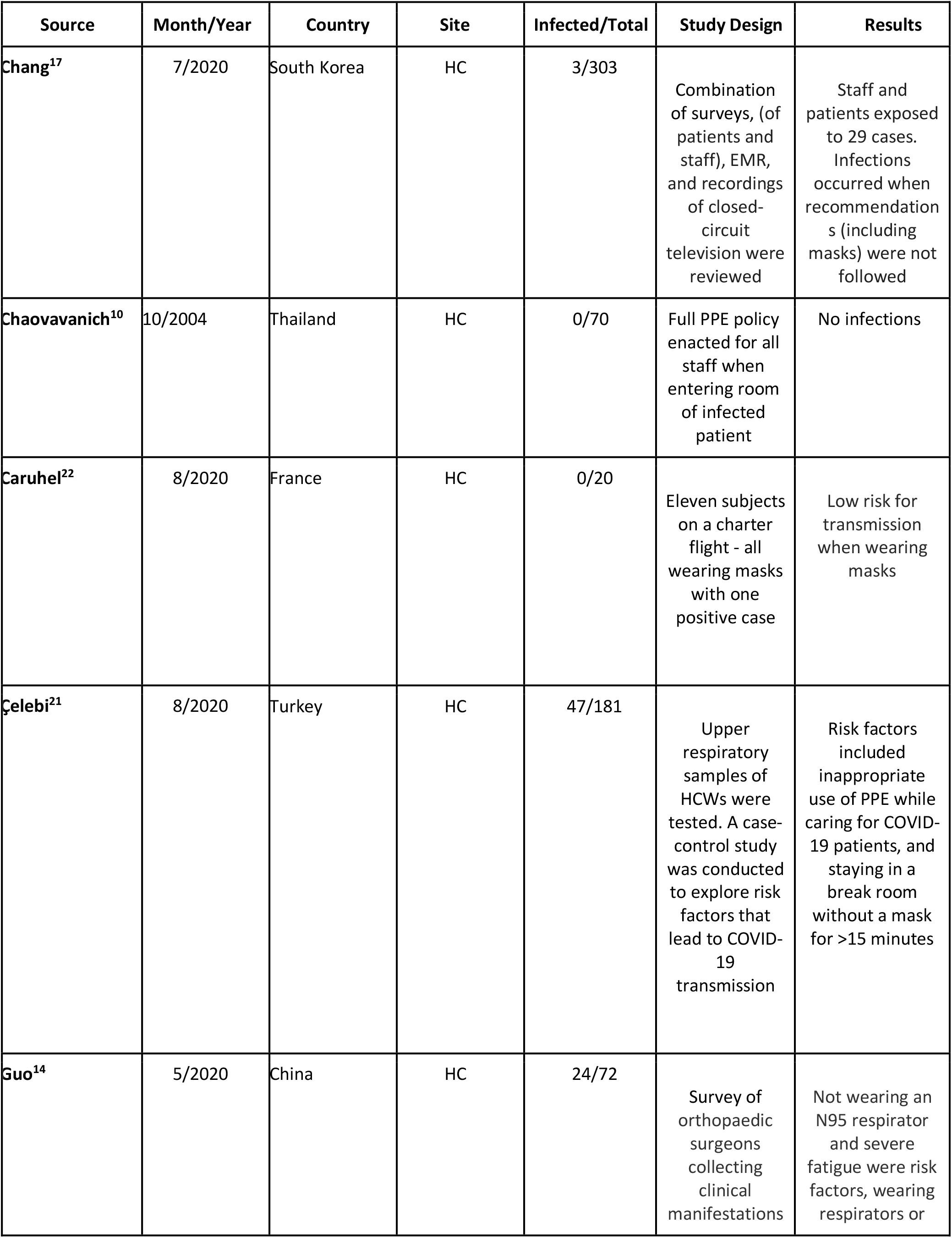
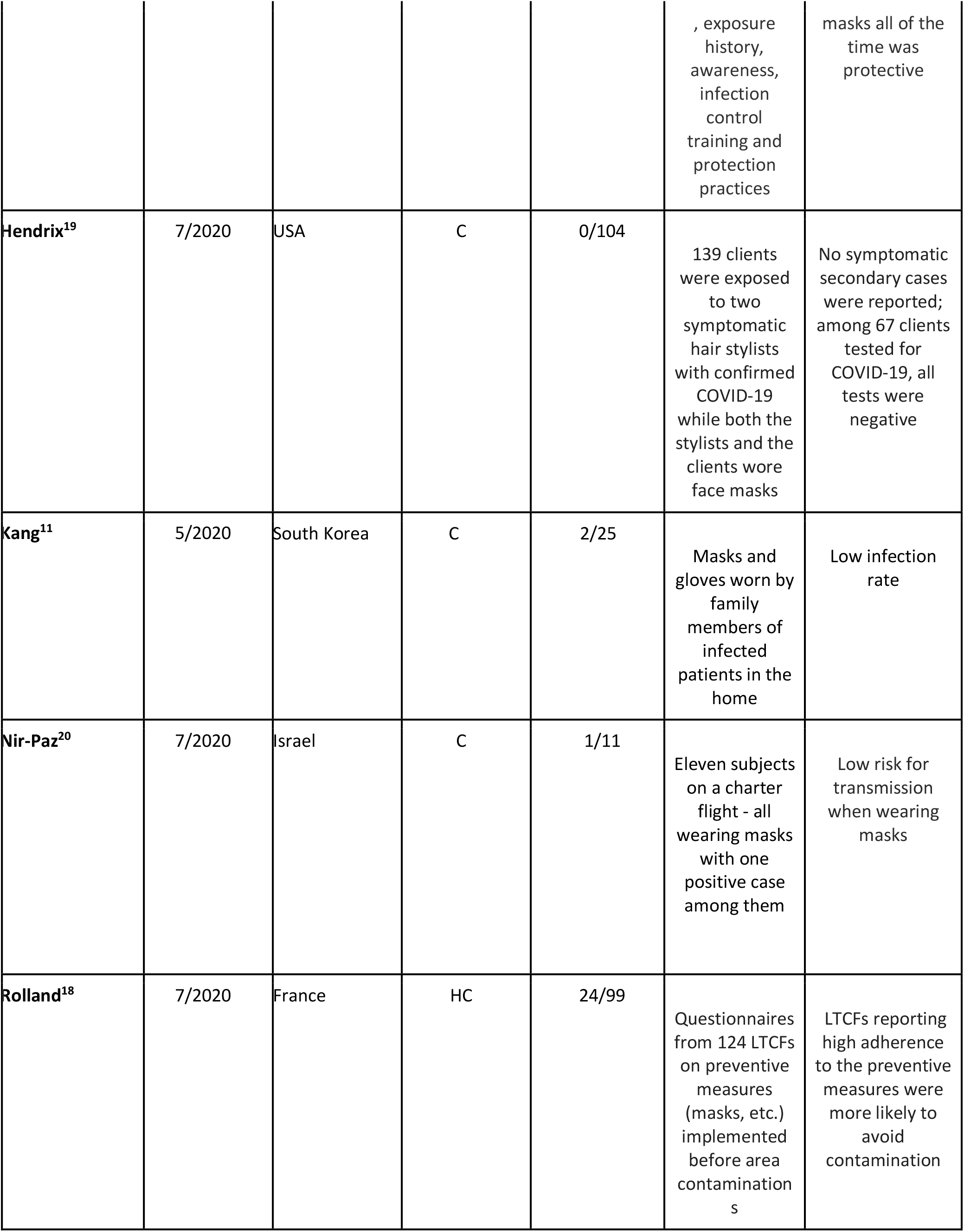
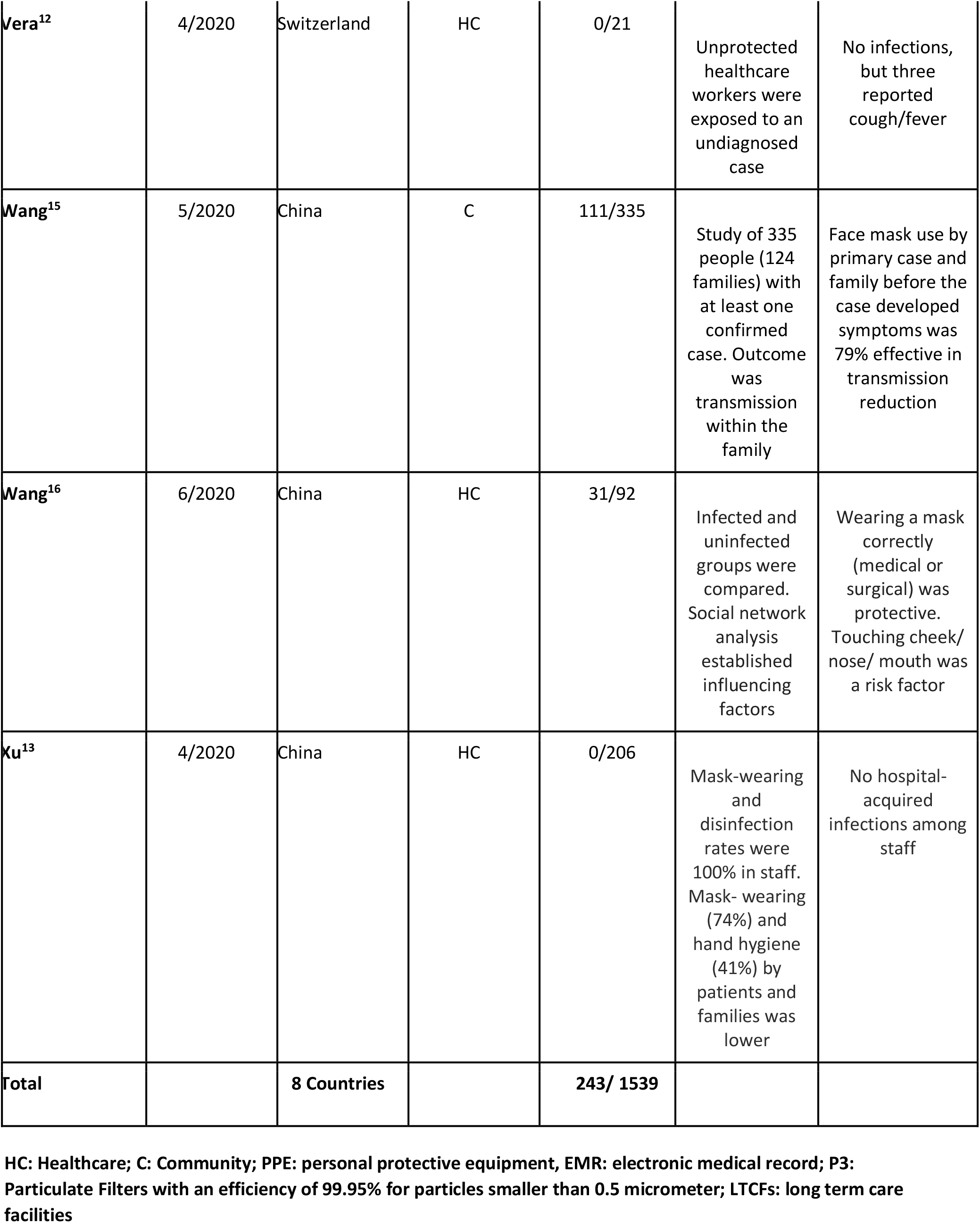
Summary of Included Studies.

*Figure 3* - For subjects in health care settings (n=1076), 74/873 (9%) of mask wearers received a positive COVID test while 67/203 (33%) of non-mask wearers received a positive test. Relative Risk of getting COVID-19 for mask wearers in the health care setting was 0.20 (95% CI: 0.15-0.27).

**Figure 3.**
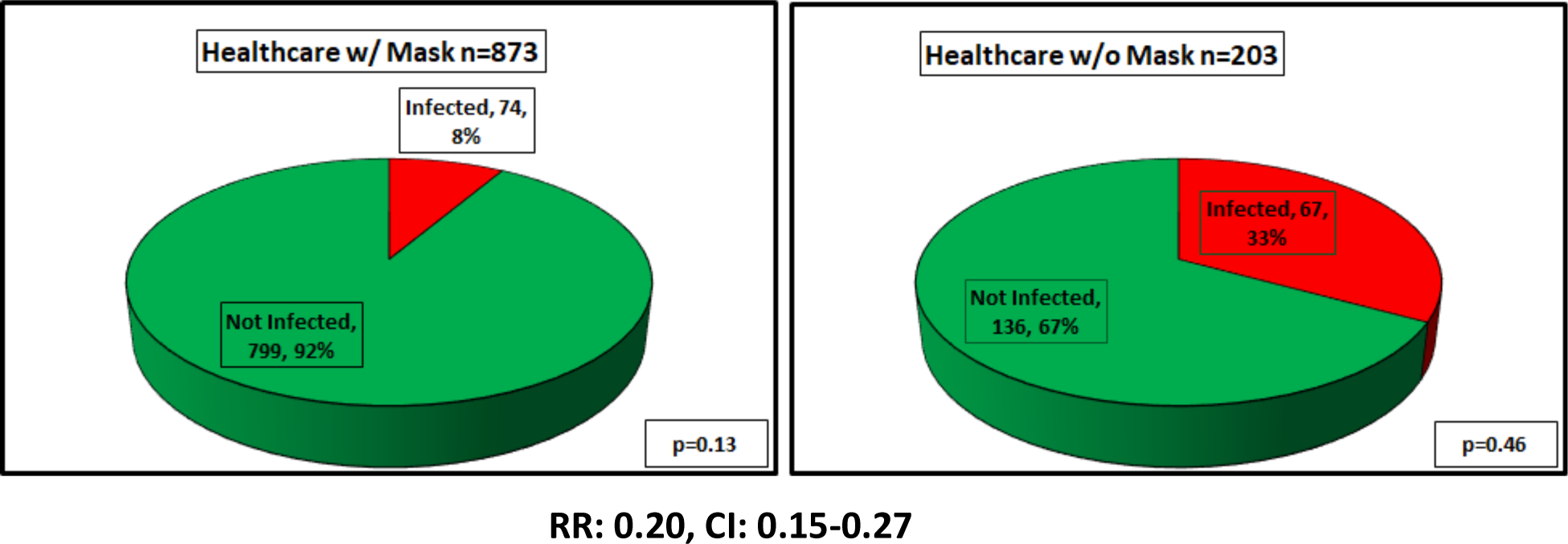
Subjects in a Healthcare Setting Infected and Not Infected with COVID-19.

*Figure 4* - In community settings (n=475), 23/365 (6%) of mask wearers received a positive COVID test while 91/110 (83%) of non-mask wearers received a positive test. Relative Risk of getting COVID-19 for mask wearers was 0.08 (95% CI: 0.05-0.11).

**Figure 4.**
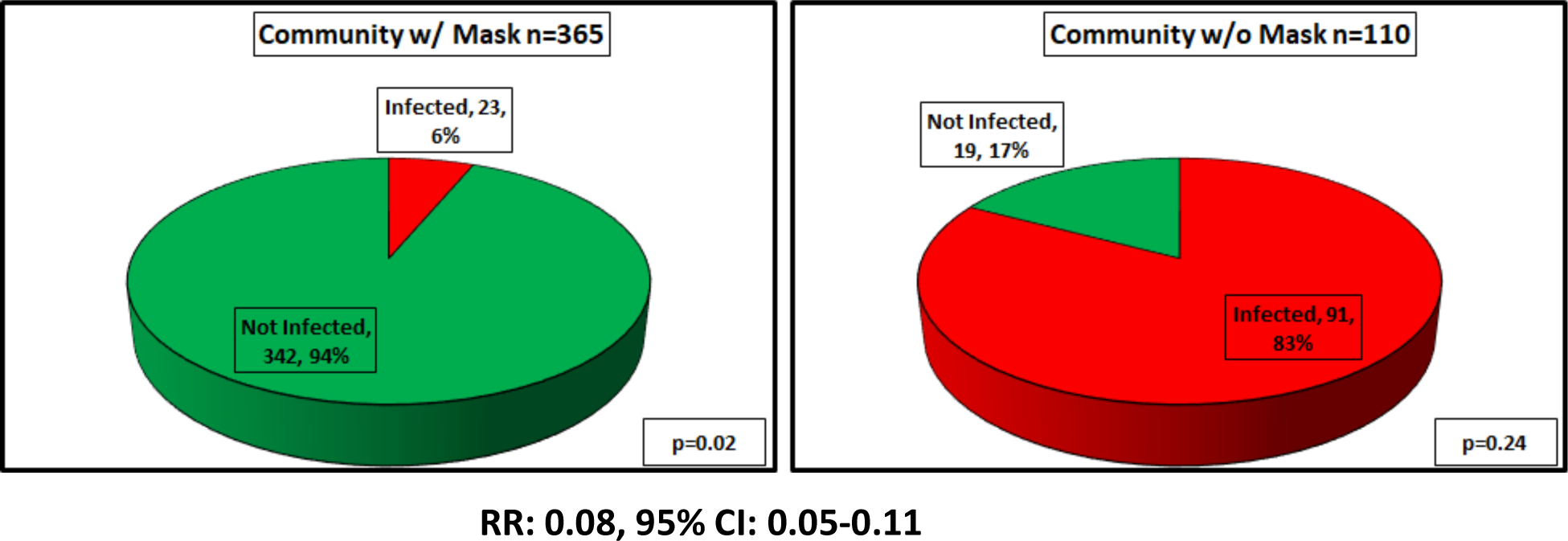
Subjects in a Community Setting Infected and Not Infected with COVID-19.

## Discussion

Based on these results, we determined that across healthcare and community settings, those who wore masks were less likely to contract COVID-19. In health care settings, a smaller percentage of individuals contracted COVID-19 than in the community (13.8 versus 30.7 percent), and of those who contracted COVID-19, a smaller percentage wore face coverings (43.9 versus 56.1 percent).

The results have shown a correlation between wearing a mask and contracting COVID-19 (regardless of healthcare or community setting) with more than 92% of people in the included studies not getting COVID-19 when they were wearing a mask (figure 2 - right graph). The correlation between not wearing a mask and contracting COVID-19 is more variable between healthcare settings and community settings. Overall, about 50% of individuals not wearing a mask in the included studies did not get COVID-19 (figure 2 - left graph). However, there was a much larger correlation with not wearing a mask and contracting COVID-19 in the community setting compared to the healthcare setting. In the community setting 83% of individuals contracted COVID-19 when not wearing a mask (figure 4 - right graph) as compared to 33% of individuals in healthcare settings (figure 3 - right graph).

One possible explanation for this disparity is that individuals in healthcare settings are more cautious with identifying and isolating infected individuals because of higher perceived risk. It is also possible that individuals in the healthcare setting may be tested more often, and thus more likely to be aware of infection before the onset of symptoms, allowing quarantine prior to exposing additional people.

Since our systematic review included studies of all types, we also considered that in the healthcare setting there is greater access to N95 masks, which are usually used in conjunction with other masks such as surgical masks or masks with face shields, all of which decreased the risk of contracting COVID-19 for healthcare workers.

Articles that were considered included case reports, cross-sectional studies, case-control studies, and randomized controlled trials. Case-control studies and randomized controlled trials did not contain human subjects but rather tested particle transmission on specific types of masks. Inclusion based on date of publication included all studies published through July 2020 with studies that were predominantly published between April 2020 and July 2020. Only peer reviewed or pre-print studies were considered.

We used this data to make an evidence-based correlation between wearing masks of any type and the incidence of COVID-19 disease or positive COVID-19 test.

Some limitations include the fact that studies differed in terms of type of mask used/tested, inability to control for all circumstances wherein an individual subject could have been exposed to COVID-19, and inability to control for proper mask usage. There are also limitations based on the cutoff period for studies considered for statistical analysis (i.e. publication through July 2020). In addition, ethical standards precluded randomized controlled trials to determine rates of COVID-19 based upon mask usage.

Facemasks are rarely used as a sole means for infection prevention. Other preventative measures, such as distancing and hand hygiene, for example are typically used in conjunction with them. A possible avenue of future research would involve analysis of which measures are most effective when used in combination with facemasks. Additionally, there is considerable variability in regional laws that govern the use of facemasks. In some areas, facemasks are simply recommended by officials; whereas, in other areas there are legal and financial penalties for not wearing a facemask. Future investigations may study the effects of such legal mandates. Finally, this initial sample size was relatively small, and additional analysis will be warranted as more information becomes available.

## Data Availability

All data produced in the present work are contained in the manuscript.

